# The association between sleep duration with migraine frequency, disability and perceived stress in the Headache Assessment via a Digital platform in United States (HeAD-US) Study

**DOI:** 10.1101/2025.10.23.25338639

**Authors:** Angeliki Vgontzas, Kristina M. Fanning, Ryan C. Bostic, Alexandre Urani, François Cadiou, Richard B. Lipton, Ali Ezzati

## Abstract

**Study objectives:** To examine the association between self-reported sleep duration on migraine frequency and disability

**Methods:** We conducted a cross-sectional analysis from the Headache Assessment via a Digital Platform in the United States (HeAD-US) Study, a survey of adult users of the Migraine Buddy App. Participants reported average nightly sleep duration, headache features and completed the Perceived Stress Scale-4, the Migraine Disability Assessment Test and the Patient Health Questionnaire-4 for anxiety and depression. Sleep duration was categorized as short (≤6 hours), normal (7-9 hours) and long (≥10 hours). Migraine diagnosis adhered to the International Classification of Headache Disorders-3.

**Results:** The 6267 participants had a mean age of 41.5±13.1 years, were 90.8% female and had a mean sleep duration of 6.92 hours (SD 1.32, range 2–15). Compared to normal sleepers, short sleepers had higher risk of more frequent monthly headache days (RR = 1.128, 95% CI: 1.067–1.192) and disability (RR = 1.167, 95% CI: 1.106–1.231). Long sleepers had greater risk of more frequent monthly headache days (RR = 1.274, 95% CI: 1.049–1.464) and disability (RR = 1.699, 95% CI: 1.446–1.997). In mediation models, short sleep was linked to approximately 1.3 extra monthly headache days and long sleep to approximately 3 extra days, with stress accounting for only part of these effects.

**Conclusions:** Unhealthy sleep duration, particularly long sleep, is associated with greater migraine burden, characterized by increased headache frequency and disability. Perceived stress partly mediated but did not fully account for these relationships.

**Statement of significance:** This study is the first to comprehensively report on associations of unhealthy sleep duration with migraine burden in a large population of adults with migraine. Adults who report sleeping too little (6 hours ore less per night) or too much (10 hours or more per night) on average have more frequent and disabling migraine. Short sleep was linked to approximately 1.3 extra monthly headache days and long sleep to 3 extra days, with stress accounting only for part of these effects.

## Background

Short and long self-reported sleep duration are associated with adverse health outcomes, including cardiovascular disease and dementia^1,2^. Evidence across large cohorts supports a U-shaped relationship between sleep duration and multiple health risks, motivating attention to both tails of the distribution (short and long sleepers).^2-5^ While optimal sleep duration varies by individual, consensus recommendations identify 7–9 hours of nightly sleep as the healthy range for adults.^6^

The associations between sleep and migraine are complex and nuanced. Meta-analytic evidence shows consistent worse subjective sleep quality in adults with migraine and suggests interictal differences in polysomnographic architecture, primarily in children.^7^ Several sleep disorders—especially insomnia and restless legs syndrome—are comorbid with migraine^7-14^.

Too little or too much sleep is a commonly reported attack trigger; however prospective daily studies do not support immediate night-to-next-day links for duration alone^15-17^. In contrast, sleep fragmentation, quality and cumulative unhealthy sleep patterns, along with stress over days or weeks may be more informative for headache risk.^15-21^

Adults with migraine have similar average nightly sleep duration compared to those without migraine (approximately 10 minutes less per night), ^7,9,21^ but the clinical impact of “unhealthy” duration at either tail is less well defined. Several clinic-based studies have reported associations between short sleep duration and greater headache frequency, typically amounting to two to three additional monthly headache days ^22,23^. However, these studies were often underpowered to detect effects of long sleep duration and did not consistently adjust for psychological comorbidities such as depression or anxiety or explicitly test stress as a mediator. Chronic stress negatively impacts sleep and is associated with worse headache frequency and severity^19,20,24^. Daily-diary and actigraphy studies suggest that multidimensional “sleep health” (regularity, timing, satisfaction, duration, etc.) may better capture migraine risk than duration alone, motivating models that incorporate perceived stress alongside sleep parameters.^21^

Stress plausibly mediates or modifies sleep–migraine associations, as adults with migraine report high perceived stress. Prior work shows interactions between sleep metrics and perceived stress in predicting headache occurrence and severity, but formal mediation has rarely been tested in large, well-phenotyped cohorts. To address this gap, we use data from the large sample from Headache Assessment via Digital Platform in the United States (HeAD-US) Study to examine whether short (≤6 h) and long (≥10 h) average nightly sleep are associated with migraine frequency, disability, and perceived stress, and whether perceived stress partially mediates these relationships after accounting for key covariates. We hypothesized that adults with migraine who report short or long average nightly sleep duration would have higher headache frequency, greater migraine-related disability, and elevated perceived stress compared to those with normal sleep (7–9 hours). We further hypothesized that perceived stress would partially mediate these associations.

## Methods

### Study design and participants

We conducted a cross-sectional analysis using baseline data from the Headache Assessment via a Digital Platform in the United States (HeAD-US) Study, a large, app-based observational study of adults with migraine. Detailed methodology has been previously reported ^25^. In brief, the HeAD-US is a cohort of adults with migraine surveyed through the Migraine Buddy smartphone app, which is commercially available and intended for individuals with headache to keep a headache diary and obtain summary reports. Several studies have been previously published using app-based data ^26-28^. In September 2023 and over three months, app users were invited to participate in the HeAD-US study. Participation involved completing a questionnaire on sociodemographic information, headache features, psychiatric symptoms and use and response to migraine-targeted treatments. Participation was voluntary and all participants provided electronic consent and were not compensated.

A total of 6810 app users completed the questionnaire, and all participants completed the AMS/AMPP migraine diagnostic module. Individuals were eligible for inclusion in the current study if they (1) self-reported a physician diagnosis of migraine, (2) met modified International Classification of Headache Disorders, 3rd edition (ICHD-3) criteria based on the American Migraine Study/American Migraine Prevalence and Prevention (AMS/AMPP) diagnostic module, (3) completed the Perceived Stress Scale-4 (PSS-4), and (4) responded to the question assessing average nightly sleep duration. Based on these criteria, 6,267 individuals were eligible for the present analysis.

### Study measures

Participants were surveyed for sociodemographic information including their age, biological sex (male or female), annual household income, highest level of education (high school degree or less, high school degree and less than 4 years of college or college degree or more). Sleep duration was self-reported in a single item: “On average, how many hours do you sleep every night?” Stress was assessed using the Perceived Stress Scale-4 total score (PSS-4) a validated measure with scores ranging from 0 to 16 ^29^. Migraine-related disability was assessed using the Migraine Disability Assessment Test (MIDAS) total score, which classifies disability as none/mild (score ≤10), moderate (11–20), or severe (≥21) ^30^. Anxiety and depressive symptoms were screened as present or absent using the Patient Health

Questionnaire - 4 (PHQ-4), with binary indicators for anxiety and depressive symptoms (present/absent) ^31^. Migraine characteristics, including self-reported number of average monthly headache days and additional headache features per ICHD-3 via the AMS/AMPP diagnostic module.

### Statistical Analysis

Descriptive statistics were used to summarize all study measures and reported as means (standard deviations) for continuous variables and frequencies (percentages) for categorical variables. Sleep duration was categorized as: short (≤6 hours), normal (7-9 hours) and long (≥10 hours). These thresholds were based on recommendations from the American Academic of Sleep Medicine and Sleep Research Society, which define 7–9 hours as the optimal range for healthy adult sleep ^32^. Throughout the manuscript we reference ≤6 hours of sleep as “short sleep duration” and ≥10 hours as “long sleep duration”.

Group differences across sleep categories were assessed using chi-square tests for categorical and one-way ANOVA for continuous variables. To examine associations between sleep duration and migraine burden, we employed multivariable negative binomial regression models with normal sleep (7–9 hours) as the reference group. The two co-primary outcomes were monthly headache days (MHD), and migraine-related disability based on the MIDAS score. Linear regression was used to assess the relationship between sleep duration and perceived stress (PSS-4, total score). All models were all adjusted for age, sex, income and education. To evaluate the additional contribution of psychological comorbidity, a second model further adjusted for anxiety and depression.

To explore whether perceived stress mediates the relationship between sleep duration and the two primary outcomes (MHD and MIDAS), we conducted a series of mediation analyses using PROCESS macro version 4.2 for SPSS (Model 4; Hayes, 2022). Separate models assessed the indirect effects of short and long sleep duration (each compared to normal sleep) on MHD and MIDAS scores via perceived stress, adjusting for age, sex, income, and education. MIDAS scores were log-transformed due to right-skewed distribution and violation of normality assumptions. Indirect effects were estimated using 5,000 bootstrap samples with bias-corrected 95% confidence intervals. We report standardized coefficients for all key mediation model components: the effect of sleep duration on perceived stress (path a), the effect of stress on each outcome (path b), the indirect effect (a×b), the direct effect of sleep duration on the outcome after adjusting for stress (path c′), and the total effect (path c). All analyses were conducted using SPSS software and all hypothesis tests were two-tailed and considered significant at p < 0.05, with test statistics and corresponding p-values reported for all comparisons.

## Results

### Sample Characteristics

A total of 6267 participants were included, with a mean age of 41.5±13.1 years and 90.8% female (Table 1). The average self-reported nightly sleep duration was 6.92 hours (SD 1.32, range 2–15), with 35.3% reporting short sleep (≤6 hours), 62.2% reporting normal sleep (7–9 hours), and 2.5% reporting long sleep (≥10 hours). Compared to those with normal sleep duration, individuals with short or long sleep duration were more likely to report lower income (<$75,000/year), lower educational level, higher migraine-related disability scores (MIDAS), and higher rates of anxiety and depressive symptoms (p <0.001 for all, Table 1). Long sleepers had a higher median migraine headache days per month (12) than normal or long sleepers (10).

**Table 1.** Sample characteristics stratified by self-reported average sleep duration.

|  | **≤6 hours** | **7-9 hours** | **≥10 hours** | **Total** |
| --- | --- | --- | --- | --- |
| N (%) | 2215 (35.3) | 3896 (62.2) | 156 (2.5) | 6267 |
| Age, Mean (SD) | 43.1 (12.7) | 40.9 (13.2) | 36.4 (14.0) | 41.5 (13.1) |
| Female, n (%) | 1952 (88.1) | 3598 (92.4) | 142 (91.0) | 5692 (90.8) |
| Race, White (%) | 1746 (78.8) | 3411 (87.6) | 142 (91.0) | 5299 (84.6) |
| Ethnicity, Hispanic (%) | 170 (7.7) | 226 (5.8) | 8 (5.1) | 404 (6.4) |
| Annual Household Income, >\$75,000, n (%) | 795 (45.1) | 1691 (51.5) | 42 (36.5) | 2528 (50.7) |
| Education, College degree or more, n (%) | 1491 (67.3) | 2972 (76.3) | 92 (59.0) | 4555 (72.7) |
| Migraine headache days per month, median (Q<sub>1</sub>, Q<sub>3</sub>) | 10 (5, 10) | 10 (5, 15) | 12 (6, 22) | 10 (5, 17) |
| MIDAS Score, median (Q<sub>1</sub>, Q<sub>3</sub>) | 59 (28, 120) | 45 (23, 93) | 76 (39, 212) | 50 (25, 105) |
| PSS-4 Total Score, mean (SD) | 8.13 (3.07) | 7.14 (3.05) | 8.98 (3.47) | 7.53 (3.11) |
| Anxiety, PHQ4, n (%) | 1099 (49.6) | 1476 (37.9) | 90 (57.7) | 2665 (42.5) |
| Depression, PHQ4, n(%) | 814 (36.7) | 989 (25.4) | 71 (45.5) | 1874 (29.9) |

Data were missing for sex (n=62), race (n=238), education (n=185), ethnicity (n=119), income (n = 1283), anxiety (n =1)

### Primary outcomes

Short and long sleep were significantly associated with higher monthly headache frequency compared to normal duration sleep. Specifically, short sleep was associated with a 12.8% increased risk of more frequent MHDs (RR = 1.128, 95% CI: 1.067,1.192), corresponding to approximately 1.3 additional headache days per month. Long sleep duration was associated with a 27.4% increased risk of chronic migraine (RR = 1.274, 95% CI: 1.049– 1.464) or about 3 additional headache days per month (Table 2). Compared to normal sleepers, individuals with short sleep had 16.7% higher risk of disability (RR = 1.167, 95% CI: 1.106– 1.231, p < 0.001), while long sleepers had 69.9% higher risk of disability (RR = 1.699, 95% CI: 1.446–1.997, p < 0.012).

**Table 2.**
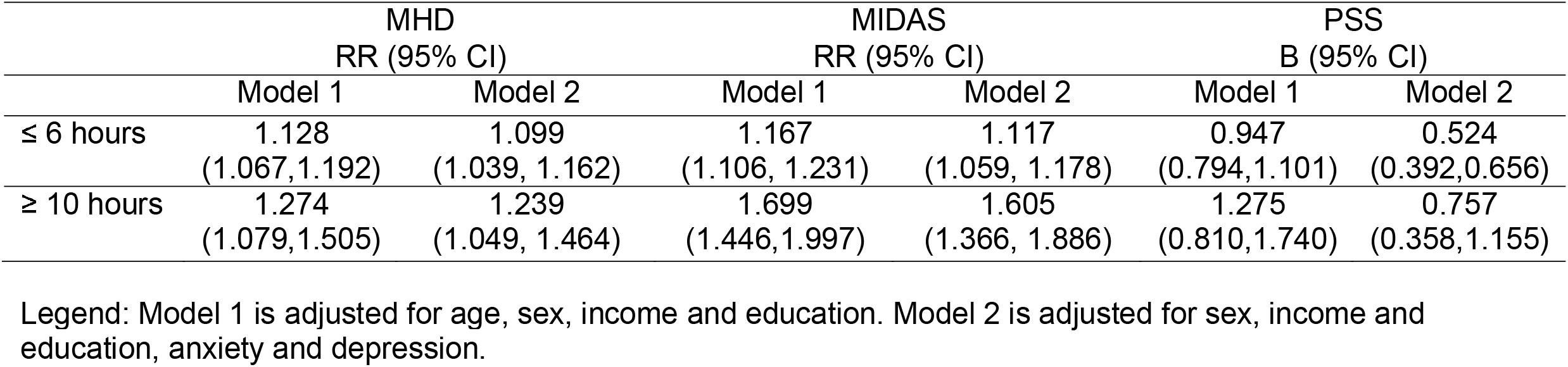
Association between abnormal sleep duration (compared to normal sleep duration) and migraine headache days per month (MHD), disability (MIDAS) and stress (PSS), adjusted models.

Short sleepers had higher perceived stress scores (B = 0.947, 95% CI: 0.794–1.101), as did long sleepers (B = 1.275, 95% CI: 0.810–1.740, p =<0.001). These associations remained significant, though slightly attenuated, after further adjustment for anxiety and depressive symptoms (Table 2). A J-shaped relationship was observed between sleep duration and perceived stress, with both short and long sleepers experiencing higher stress levels, especially among individuals with chronic migraine (Figure 1).

**Figure 1.**
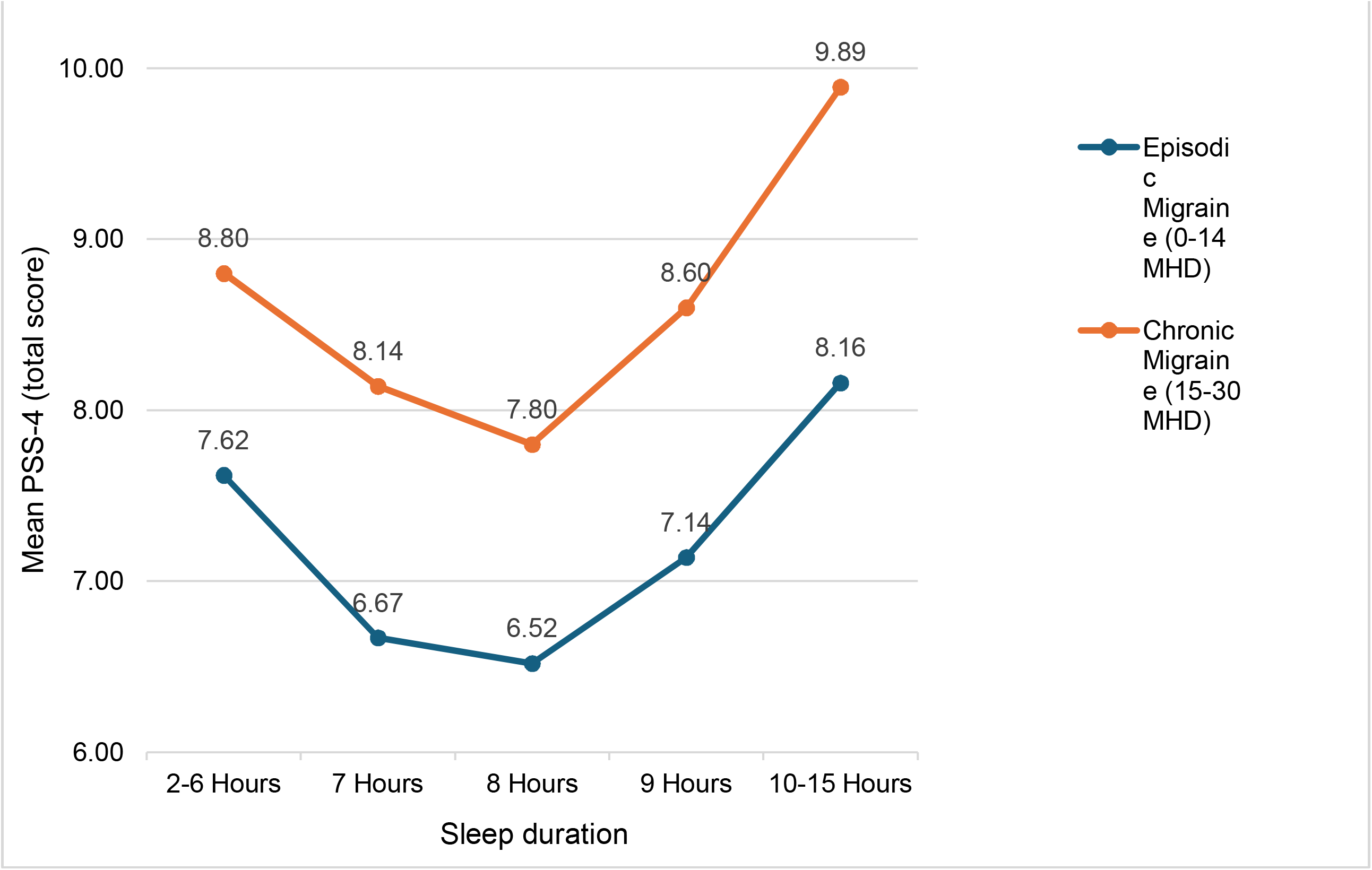
Sleep Duration and Perceived Stress in the HeAD-US Study.

### Mediation models

We investigated whether perceived stress mediated the association between sleep duration and migraine burden on MHD outcome, adjusting for age, sex, income and education (Figure 2). For MHD, short sleep duration showed both direct (β = 0.0756, 95%CI: 0.001 – 0.150) and indirect effects via stress (β = 0.0826, 95%CI: 0.066 – 0.100; Figure 2a). This corresponded to an estimated 1.3 additional MHDs among short sleepers compared to normal sleepers, of which approximately 0.62 days were attributable to non-stress pathways. Similarly, long sleep duration was also associated with a significant direct effect on MHD (β= 0.280, 95%CI: 0.003 – 0.557) and an indirect effect via stress (β= 0.085, 95%CI: 0.033, 0.138), corresponding to roughly 3 more MHDs, with 2.3 days remaining after accounting for perceived stress (Figure 2a). Overall, the magnitude of these associations was stronger for long duration sleepers than for short duration sleepers when compared to normal duration sleepers.

**Figure 2.**
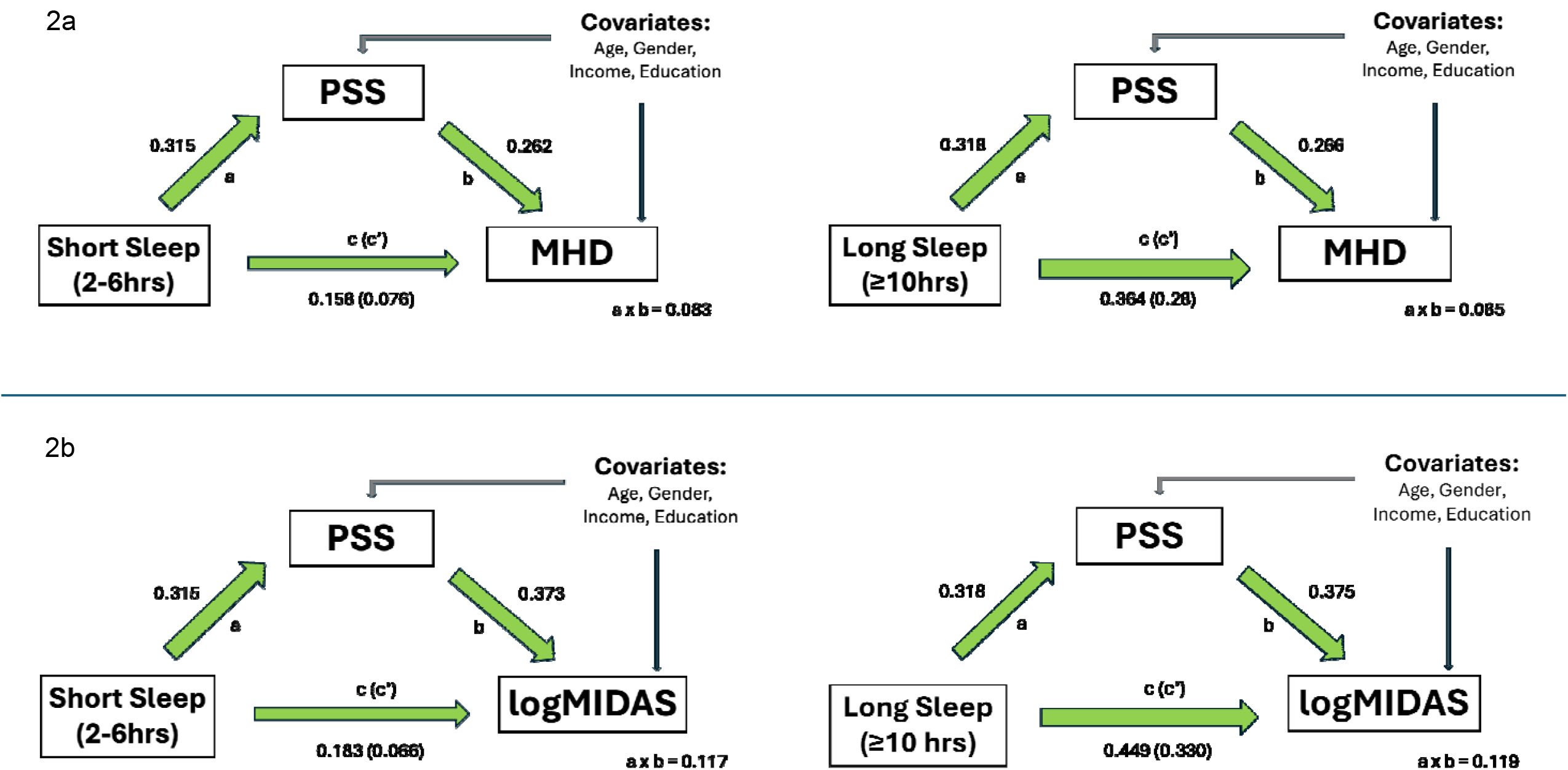

We also examined the mediating role of perceived stress in the relationship between sleep duration and migraine-related disability, measured using the log-transformed MIDAS score. For short sleep, the total effect on logMIDAS was β = 0.183 (95% CI: 0.141 – 0.225), comprising a direct effect of β = 0.066 (95% CI: 0.009 – 0.123) and an indirect effect via stress of β = 0.177 (95% CI: 0.095 – 0.140). For long sleep, the total effect was notably larger (β = 0.449, 95% CI: 0.364 – 0.534), with a direct effect of β = 0.330 (95% CI: 0.250 – 0.409) and an indirect effect of β = 0.119 (95% CI: 0.050 – 0.188). These estimates suggest that long sleep duration was associated with a 23.2% increase in MIDAS score (16.6% attributable to direct effects), while short sleep was associated with a 6.1% increase (3.0% directly). The inclusion of anxiety and depression as additional covariates did not substantially alter these mediation effects for both MHD and logMIDAS.

## Discussion

In this large, app-based cohort of adults with migraine, both short (≤6 hours) or long (≥10 hours) average nightly sleep duration were associated with greater migraine burden, including a more headache days per month, more severe disability, and higher perceived stress. The magnitude of these associations was strongest for long sleepers, who experienced an additional three days of migraine per month. Perceived stress partially mediated, but did not fully account for, these relationships, and the results remained robust after adjustment for co-occurring anxiety and depression. Taken together, these findings suggest that self-reported sleep duration is a clinically meaningful indicator of migraine severity, with long sleep identifying a subgroup at particularly elevated risk of disability.

Both short and long sleepers showed more unfavorable comorbidities and sociodemographic profiles compared to normal sleepers, consistent with greater migraine severity (see Table 1). Long sleepers, despite being on average younger (36.4 years vs. 40.9– 43.1 years), had the highest rates of anxiety, depression symptoms, and perceived stress, as well as lower levels of income and educational attainment. These findings are largely consistent with known associations between short and long sleep with less advantageous sociodemographic characteristics literature and risk of depression.^33-35^ Although sleep duration changes across the lifespan, it remains relatively consistent in middle age (35-50 years) which suggests that long sleep may represent a distinct clinical subgroup, characterized by greater disability emerging earlier in adulthood.^36^

We report that both short and long sleep are associated with worse migraine frequency and disability. Few studies have previously reported on average sleep duration in cohorts of patients with migraine ^21-23^. Consistent with our findings, self-reported short sleep duration (<6 hours) and long sleep duration (>8 hours) have been associated with worse headache frequency in clinical samples of adults with migraine, with an additional 2-3 headache days per month for short sleepers compared to healthy sleepers. However, these studies were underpowered to examine long sleep and did not adjust for psychiatric comorbidity or perceived stress ^22,23^. One smaller study of episodic migraine found no direct association between sleep duration and headache frequency but identified sleep duration as a contributor to a multidimensional sleep health score in which healthier sleep predicted three to four fewer monthly headache days^21^. By analyzing more than 6,000 participants, over one-third with chronic migraine, we confirmed a J-shaped association: both short and long sleep predict greater headache frequency and disability, with the strongest effects in long sleepers (see Figure 1). Importantly, findings were unchanged after adjustment for anxiety and depressive symptoms, underscoring the independent role of sleep duration.

We found that perceived stress acted as an indirect mediator of the associations between short and long sleep and greater migraine burden; however, the direct effects of sleep were stronger. Long sleepers had ∼3 additional MHDs, of which ∼2.3 persisted after accounting for stress. The analysis examining migraine-related disability showed a similar pattern, most of which reflected direct rather than stress-mediated pathways. Prior studies report that changes in perceived stress precede headache days and interact with sleep duration to influence headache severity in those with chronic migraine ^19,24^. Evidence for more chronic interactions is limited, but in adults with episodic migraine, baseline reports of poor sleep quality predicted higher recurrence of headache attacks over the next six weeks, particularly among those with moderate/high perceived stress ^20,21^. Taken together, these analyses emphasize that unhealthy sleep duration is directly associated with migraine frequency and disability and to a less extent, indirectly through higher perceived stress.

Average nightly sleep duration appears to influence migraine burden both directly and indirectly through stress pathways. Prospective research suggests that these associations cannot be attributed solely to migraine attacks, indicating deeper mechanistic links between sleep and migraine susceptibility ^15-17^. These links may reflect distinct migraine endophenotypes such that short sleep may signal insomnia or hyperarousal phenotypes. Alternatively, they may arise from a bidirectional cycle in which worsening migraine severity disrupts sleep and poor sleep exacerbates migraine burden. Animal studies report that sleep deprivation lowers the threshold for propagating a migraine attack in chronic migraine models and that pro-inflammatory pathways are activated to maintain central sensitization of pain^37,38^. Insomnia frequently co-occurs with migraine and may be associated with altered sleep architecture, including reductions in sleep efficiency and slow-wave sleep ^39,40^. Moreover, neuroimaging and biochemical studies show that sleep and migraine share common central nervous system circuits (including the hypothalamus) and neurotransmitters, such as serotonin, melatonin, orexins, and dopamine, suggesting overlapping pathophysiology ^41-43^. Clinically, insomnia has been associated with elevated cardiovascular risk, hyperarousal, and HPA⍰axis dysregulation, which is a profile that could magnify migraine-related stress reactivity or contribute to the propagation of a migraine attack^44,45^. Conversely, long sleep duration in adults with migraine may stem from behavioral adaptations to high disease burden (such as extended bedrest or napping) or serve as a compensatory response aimed at metabolic restoration and glymphatic clearance post-attack^46-49^. These maladaptive patterns may perpetuate poor sleep hygiene and reinforce altered sleep–wake cycles, further compounding migraine burden.

Together, our data argue against “one-size-fits-all” sleep recommendations in migraine.

Rather, interventions may need to be tailored by sleep phenotype. For example, a subset of those reporting long sleep duration may have hypersomnia and narcolepsy which would require pharmacologic and targeted behavioral strategies for these disorder. Another subset of long duration sleepers without comorbid sleep disorders may benefit from individualized sleep hygiene recommendations and psychotherapy as part of a comprehensive strategy to address living with chronic pain. For those with short sleep duration, insomnia should be screened and treated using evidence-based therapies, such as cognitive behavioral therapy for insomnia, which also has a positive influence in reducing migraine headache days ^50^. Future longitudinal studies should evaluate temporal dynamics, incorporate actigraphy or polysomnography, and test targeted interventions.

This study has several strengths, including a large, well-characterized sample of U.S. adults with migraine, use of standardized diagnostic criteria, and validated measures of disability, stress, and psychiatric symptoms. However, several limitations should be noted. First, we lacked information on sleep disorders, such as insomnia and excessive daytime sleepiness, which may be associated with sleep duration and migraine outcomes. Second, the cross-sectional design precludes conclusions about causality or temporal direction. Third, the digitally recruited cohort underrepresents certain populations, such as older adults, individuals with limited internet access, and those from lower socioeconomic backgrounds, which may limit generalizability. Finally, recruitment through an app-based survey raises the possibility of selection bias, as individuals who chose to participate may differ systematically from those who did not.^25^

Self-reported short and long sleep duration is associated with a more disabling and more severe migraine phenotype, characterized by increased headache frequency, greater symptom severity, and higher stress levels. Further investigation is needed to characterize the sleep patterns and symptoms in this group of patients to inform individualized interventions.

## Data availability

Data supporting the findings of this study are available from the study team upon reasonable request.

